# The role of accelerometer-derived sleep traits on glycated haemoglobin and glucose levels: a Mendelian randomization study

**DOI:** 10.1101/2022.10.11.22280427

**Authors:** Junxi Liu, Rebecca C Richmond, Emma L Anderson, Jack Bowden, Ciarrah-Jane S Barry, Hassan S Dashti, Iyas S Daghlas, Jacqueline M Lane, Simon D Kyle, Céline Vetter, Claire L Morrison, Samuel E Jones, Andrew R Wood, Timothy M Frayling, Alison K Wright, Matthew J Carr, Simon G Anderson, Richard A Emsley, David W Ray, Michael N Weedon, Richa Saxena, Martin K Rutter, Deborah A Lawlor

**Author notes:** Joint first authors, with equal contributions. Joint senior authors, with equal contributions (Martin K Rutter, Deborah A Lawlor). Corresponding author: Dr. Junxi Liu, MRC Integrative Epidemiology Unit, Bristol Medical School, University of Bristol Oakfield House, Oakfield Grove, Clifton, Bristol, BS8 2BN.

## Abstract

**Study Objectives:** Self-reported shorter/longer sleep duration, insomnia, and evening preference are associated with hyperglycaemia in observational analyses, with similar results observed in small studies using accelerometer-derived sleep traits. Mendelian randomization (MR) studies support an effect of self-reported insomnia, but not other sleep traits, on glycated haemoglobin (HbA1c). Our aims were a) to explore potential effects of accelerometer-derived sleep traits on HbA1c and glucose levels and b) to determine genetic correlations across accelerometer-derived and self-reported sleep traits.

**Methods:** We used MR methods to explore effects of accelerometer-derived sleep traits (duration, mid-point least active 5-hours, mid-point most active 10-hours, sleep fragmentation, and efficiency) on HbA1c in European adults from the UK Biobank (UKB) (n = 73,797) and the MAGIC consortium (n = 149,054). Cross-trait linkage disequilibrium score regression was also applied to determine genetic correlations across all accelerometer-derived and self-reported sleep traits and HbA1c/glucose.

**Results:** Main and sensitivity MR analyses showed no causal effect of any accelerometer-derived sleep trait on HbA1c or glucose. Similar MR results for self-reported sleep traits in the UKB sub-sample with accelerometer-derived measures suggested our results were not explained by selection bias. Genetic correlation analyses suggested complex relationships between self-reported and accelerometer-derived traits indicating that they may reflect different types of exposure.

**Conclusions:** Taken together, these findings suggested accelerometer-derived sleep traits do not causally affect HbA1c levels, and accelerometer-derived measures of sleep duration and sleep quality might not simply be ‘objective’ measures of self-reported sleep duration and insomnia, but rather captured different underlying sleep characteristics.

**Statement of Significance:** Self-reported and accelerometer-derived sleep disturbance is associated with increased risk of hyperglycaemia and type 2 diabetes in observational analyses. Mendelian randomization (MR) studies support an effect of self-reported insomnia, but not other self-reported sleep traits, on glycated haemoglobin (HbA1c). This MR study showed little evidence supporting an effect of any accelerometer-derived sleep trait on HbA1c or glucose, but a potential non-linear (e.g., U-shaped) effect cannot be ruled out. The genetic correlation suggested complex relationships between self-reported and accelerometer-derived traits indicating that they may reflect different exposures.

## Introduction

Prospective cohort studies have identified associations of self-reported short and long sleep duration, insomnia (difficulty initiating or maintaining sleep), and chronotype (evening preference) with higher risks of type 2 diabetes (T2D),(1–3) hyperglycaemia and insulin resistance.(4) A small number of studies have assessed sleep characteristics using accelerometry devices, assuming these reflect similar sleep characteristics measured with greater precision and less measurement error than self-reported traits. Several observational studies showed that accelerometer-derived shorter sleep duration and lower sleep efficiency (an assumed indicator of insomnia(5)) were associated with higher glycated haemoglobin (HbA1c) levels in people with diabetes.(6, 7) In a general population, higher sleep fragmentation(8) (another indicator of insomnia(9)), but not shorter accelerometer-derived sleep duration,(10) was associated with higher HbA1c and glucose levels. However, these were relatively small studies that included ∼170(6) to ∼2107(10) participants, which are also open to residual confounding and/or reverse causation. A meta-analysis of randomized controlled trials (RCTs) showed that sleep restriction had detrimental effects on insulin sensitivity,(11) as well as hyperglycaemia supported by experimental data in healthy volunteers.(12) But the relevance of experimental sleep restriction protocols to the sleep patterns experienced in the general population is unclear.

Mendelian randomization (MR) is increasingly used to explore lifelong effects because it is less prone to confounding by social, environmental, and behavioural factors.(13) Previous MR studies showed that self-reported frequent insomnia symptoms causes higher HbA1c,(14–16) whilst no evidence has been provided for effects of self-reported sleep duration or chronotype on T2D and/or glycaemic traits.(14, 17) Recent MR studies suggested causal effects of accelerometer-derived shorter sleep duration and lower efficiency on higher waist-hip ratio but not T2D or other hyperglycaemic outcomes in the UK Biobank.(17)(18) Our aim was to explore potential effects of accelerometer-derived sleep traits (duration, mid-point least active 5-hours (L5 timing), mid-point most active 10-hours (M10 timing), sleep fragmentation, and sleep efficiency) on HbA1c. We undertook one-sample MR (1SMR) analyses using the UKB sub-sample (n = 73,797) with valid accelerometer measures. Since those with accelerometer data were not a random sub-sample of UKB, we explored possible selection bias by re-running, in this sub-sample, all of our previous MR analyses of self-reported sleep traits (duration, chronotype, insomnia) with HbA1c that had been conducted in the larger UKB sample (n = 336,999).(14) Additionally, we conducted two-sample MR (2SMR) analyses using summary outcome data from UKB and the Meta-Analyses of Glucose and Insulin-related traits Consortium (MAGIC).(19) Lastly, to help understand any differences we observed between self-reported and accelerometer-derived MR effects for assumed equivalent traits, we used cross-trait linkage disequilibrium score regression (LDSC)(20) to determine genetic correlations across all accelerometer-derived and self-reported sleep traits. We repeated all analyses with glucose as a secondary outcome.

## Methods

### The UK Biobank

Between 2006 and 2010, the UKB recruited 503,317 adults (aged 40-69 years) out of 9.2 million invited eligible adults (5.5% response).(21) Information on socio-demographic characteristics and lifestyle including self-reported sleep traits were obtained using a touchscreen questionnaire at the baseline assessment. Venous blood samples were collected and processed at baseline. A triaxial accelerometer device (Axivity AX3) was worn continuously for up to seven days in a sub-sample of participants (n = 103,711) an average of five years after the baseline assessment (range 2.8 to 8.7 years).(18) **Figure 1** shows the flowchart of participants from all recruited to those included in our study. After applying pre-specified exclusion criteria, we included 73,797 European participants(22) with accelerometer-derived sleep data in the analyses. Full details are presented in **Supplementary Information**.

**Figure 1.**
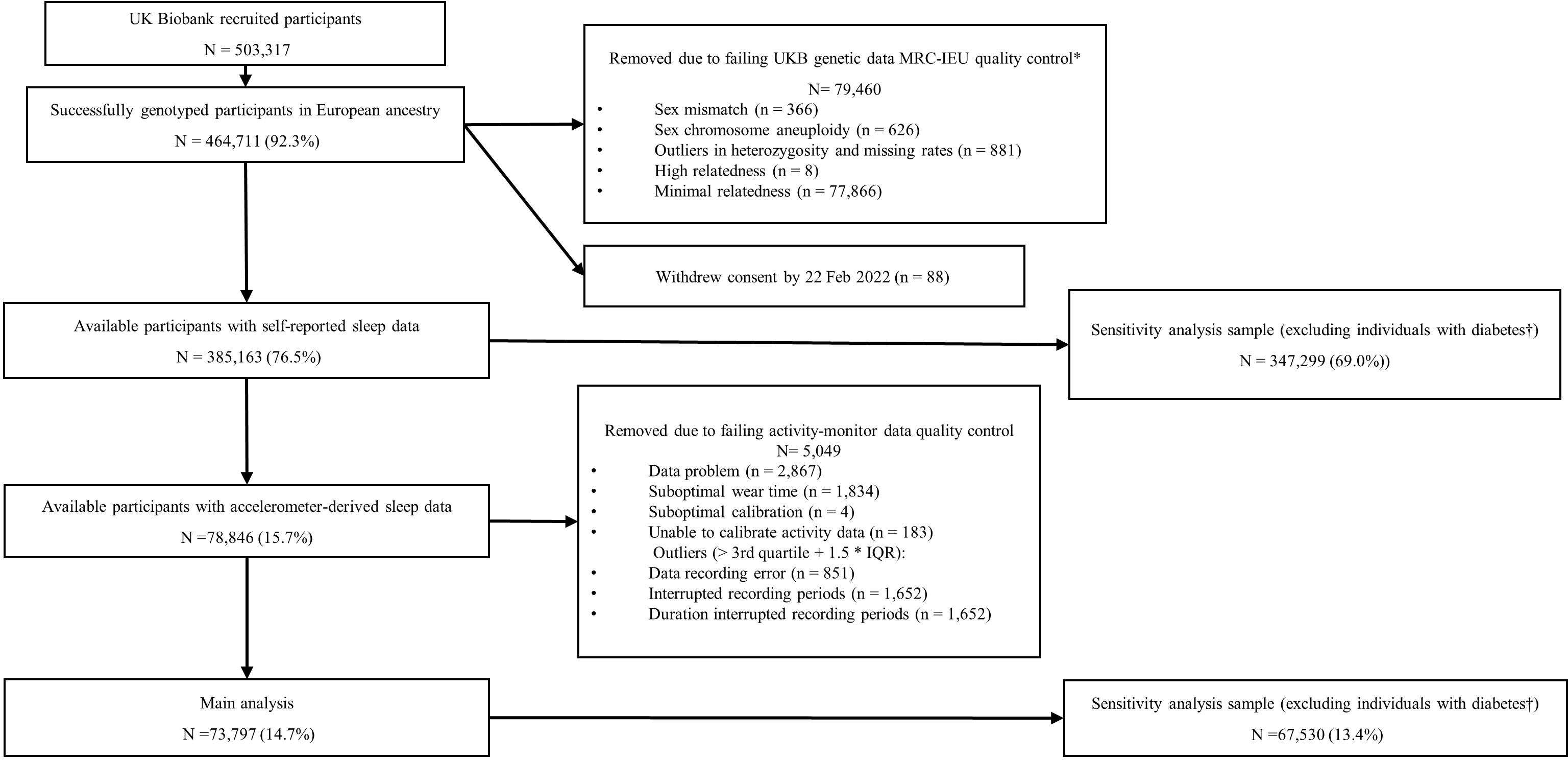
Flowchart of the participants included in the main analyses in the UK Biobank. * Quality control procedure undertaken, and the derived files produced by the MRC-IEU (University of Bristol), using the full UK Biobank genome wide SNP data (version 3, March 2018)https://data.bris.ac.uk/data/dataset/1ovaau5sxunp2cv8rcy88688v. The number of 79,460 was obtained after accounting for overlapped samples. † Excluding participants with diabetes defined by the Eastwood algorithm (probable/possible type 1 diabetes and type 2 diabetes) and/or additionally those with a baseline HbA1c ≥ 48 mmol/mol.

### Accelerometer-derived sleep traits

1. **Accelerometer-derived nocturnal sleep duration** was defined as the summed duration of all nocturnal sleep episodes within the sleep period time windows (SPT-windows). Sleep episodes were defined as any period of at least 5 minutes with no change larger than 5° associated with the z-axis of the accelerometer.(23) The algorithm in GGIR (R package) combined all sleep episodes that were not separated by more than 30 minutes and then called that the SPT-window (of which there can only be one per day). Any sleep episodes outside of this window were classified as naps and so didn’t count towards the nocturnal sleep duration total. The total duration of all SPT-windows over the activity-monitor wear time was averaged and divided by the number of days (24 hours) to give mean sleep duration per total day. Individuals with an average sleep duration < 3 (n = 147) or >12 hours (n = 3) were set to missing in this study.
2. **Midpoint least-active 5-hour (L5) timing** was a measure of the midpoint of the least-active (i.e., with minimum average acceleration) 5 hours of each day. The 5-hour periods were defined on a rolling basis (e.g., 1:00 to 6:00, 2:00 to 7:00 and so on). For example, if the midpoint of the least-active 5-hour was 24:00 (0:00) (i.e., a rolling 5-hour was from 21:30 to 2:30) then L5 = 24 (i.e., 24 + 0); if the midpoint of least-active 5-hour was 3:30 then L5 = 27.5 (i.e., 24 + 3.5); and if the midpoint of the least-active 5 hour was 20:30 then L5 = 20.5 (i.e., 24 – 3.5). Thus, a higher L5 score indicated someone was least active in the morning and more likely to have an evening chronotype.
3. **Midpoint most-active 10-hour (M10) timing** was a measure of the midpoint of the most active (i.e., with maximum average acceleration) 10-hour time of day based on a 24-hour clock. It was calculated in a similar way to L5 (see above) except with rolling periods of 10 hours. A higher M10 score indicated someone who was most active in the evening and hence more likely to have an evening chronotype
4. **Nocturnal sleep episode** (defined above) was a measure of sleep fragmentation. Individuals with an average number of sleep episodes ≤ 5 (n = 84) or ≥ 30 (n = 52) times were set to missing in this study. We referred to a high number of sleep episodes as ‘sleep fragmentation’ throughout this paper.
5. **Mean sleep efficiency** was calculated as the nocturnal sleep duration (defined above) divided by the time elapsed between the start of the first inactivity bout and the end of the last inactivity bout (which equals the SPT-window duration) across all valid nights. This was an approximate measure of the proportion of time spent asleep while in bed.

### Genetic variants

The genetic variants associated with the five accelerometer-derived sleep traits were obtained from a genome-wide association study (GWAS) conducted in UKB subsample (n = 85,670, White European), where 44 single nucleotide polymorphisms (SNPs) associated at genome-wide significance (p < 5 x 10^-8^) with at least one of the five accelerometer-derived traits (11 for sleep duration, 6 for L5 timing, 1 for M10 timing, 21 for sleep fragmentation, and 5 for sleep efficiency).(18) The genetic associations were obtained using a linear mixed model adjusting for the effects of population structure, individual relatedness, age at accelerometer assessment, sex, study centre, season of accelerometer wear, and genotype array (**Supplementary Information**).(18) **Supplementary Table S1** provides the list of SNPs used as instrumental variables for each of the accelerometer-derived sleep traits. The number of SNPs used for each accelerometer-derived sleep trait, the mean F-statistic, and variance (R^2^) across all SNPs, as well as the unweighted allele score, for each exposure are provided in **Supplementary Table S2**.

### HbA1c and glucose measurement

HbA1c was measured in red blood cells by HPLC on a Bio-Rad VARIANT II Turbo analyzer and glucose was assayed in serum by hexokinase analysis on a Beckman Coulter AU5800.(24) Samples were assumed to be non-fasting, because participants were not advised to fast before attending. The dilution factor and fasting time were considered in corresponding analyses. The HbA1c samples were not affected. We used HbA1c (a stable measure over a period of ∼ four weeks) as our primary outcome and we explored non-fasting glucose as a secondary outcome (**Supplementary Information**).

Sex-combined meta-analysis summary statistics of genetic variants related to HbA1c (%, n= ∼149,054, mean age 59.7 years, 57.9% female)) and BMI adjusted fasting glucose (mmol/l, n= ∼ 196,575, mean age 50.9 years, 51.2% female)) were also obtained from the MAGIC consortium.(19) Participants were of European descent without diagnosed diabetes.

### Statistical analyses

UKB HbA1c/glucose data were right skewed and the units (HbA1c: in mmol/mol and non-fasting glucose: in mmol/l) differed to those obtained from MAGIC(19) (HbA1c: in % and fasting glucose with BMI adjusted: in mmol/l). Therefore, we natural log-transformed the HbA1c/glucose levels in UKB and then converted them into standard deviation (SD) units (HbA1c: 1 SD = 0.14 log mmol/mol; non-fasting glucose: 1SD = 0.16 log mmol/l), as well as those from MAGIC(19) (HbA1c: 1SD = 0.41%; fasting-glucose: 1SD = 0.84 mmol/l). As such, we estimated the difference in mean HbA1c/glucose in SD units per unit increase in each accelerometer-derived sleep trait in all analyses.

Main analyses assessing the effects of accelerometer-derived sleep traits on HbA1c/glucose

#### 1SMR

We generated unweighted allele scores(25) for the sleep traits as the total number of increasing alleles present identified in the relevant GWAS(18) for each participant. Two-stage least squares instrumental variable analyses were performed to obtain the MR estimate of each trait on HbA1c/glucose. We adjusted for assessment centre and 40 genetic principal components to minimize confounding by population stratification,(26) as well as baseline age, sex, genotyping chip, fasting time and dilution factor (for glucose only) to reduce random variation.

#### 2SMR

We used summary associations between the genetic instruments and accelerometer-derived sleep traits identified in the GWAS(18) for Sample 1 (the SNP-exposure association). For sample 2 (the SNP-outcome association), we used two independent samples: **Sample 2-UKB**: estimates of the associations between the genetic instruments and HbA1c/glucose were from the sample of the UKB participants who did not participate in the accelerometer GWAS(18) (HbA1c: n = ∼292,000 and glucose: n = ∼267,000). The SNP – outcome associations were obtained via the multivariable adjusted linear model accounting for assessment centre and 40 genetic principal components, baseline age, sex, and genotyping chip, fasting time and dilution factor (for glucose only); **Sample 2-MAGIC**: the summary statistics were from the MAGIC consortium.(19) We conducted inverse-variance weighted (IVW) regression of the Wald ratio for each SNP under a multiplicative random-effects model(27) to obtain the causal estimates. Further details are presented in the **Supplementary Information**.

1SMR and 2SMR analyses taking self-reported sleep traits (sleep duration, chronotype, insomnia symptoms) as the exposures were conducted for comparison. The detailed information are presented in the **Supplementary Information**.

### Sensitivity and additional analyses

#### Accounting for the impact of diabetes

To account for the potential impact of either diabetes or the diabetic treatment on glycaemic levels, we repeated the analyses with UKB participants (1SMR and 2SMR-UKB) excluding those with diabetes defined by the *Eastwood* algorithm (probable/possible type 1 diabetes and type 2 diabetes, based on self-reported medical history and medication)(28) and/or additionally those with a baseline HbA1c ≥ 48 mmol/mol (≥ 6.5%, the threshold for diagnosing diabetes).

#### Assessing MR assumptions and evaluating bias

MR analysis requires three key assumptions to be satisfied in order to obtain valid causal estimates.(29) First, the genetic instrument should be robustly associated with the exposure. We investigated this using first-stage F-statistic and R^2^. Further details were presented in the **Supplementary Information**. An F-statistic < 10 is usually considered as a weak instrument, which may introduce weak instrument bias.(30) Second, there should be no confounding between the genetic instrument and the outcome. This can occur as a result of population stratification. We attempted to minimise this by restricting analyses to European ancestry and adjusted for genetic principal components and assessment centre.(26) Third, the genetic instrument should influence the outcome exclusively through its effect on the exposure. This would be violated by unbalanced horizontal pleiotropy (i.e., an independent pathway between the instrument genetic variant and outcome other than through the exposure). We have undertaken the following sensitivity analyses to explore potential bias due to horizontal pleiotropy.

In 1SMR, we explored between SNP heterogeneity, potentially due to horizontal pleiotropy, via the *Sargan* over-identification test.(31) Additionally, we applied the Collider-Correction(32) method to implement three further pleiotropy sensitivity analyses commonly used in 2SMR (i.e., IVW, MR-Egger, and least absolute deviation regression (LADreg) being similar to the weighted median (WM) approach). Collider-Correction was needed in 1SMR to account jointly for pleiotropy and weak instruments bias(30) (**Supplementary Information**). We subsequently referred to this as 1SMR with Collider-Correction as 1SMR-CC (i.e., 1SMR-CC-IVW, 1SMR-CC-MR-Egger, 1SMR-CC-LADreg). In 2SMR, we explored unbalanced horizontal pleiotropy by comparing the results of the IVW regression with standard pleiotropy-robust MR methods: WM and MR-Egger, referred to as 2SMR-UKB/MAGIC WM and 2SMR-UKB/MAGIC MR-Egger. To account for weak instrument bias in the 2SMR MR-Egger estimates, we used simulation extrapolation SiMEX.(33) We referred it as 2SMR-UKB/MAGIC MR-Egger_SiMEX.

#### Exploring selection bias

We compared distributions of HbA1c, glucose, diabetes prevalence, BMI, and a range of socioeconomic and behavioral characteristics (**Supplementary Information**) between those included in the sub-sample of UKB with accelerometer-derived data (n = 73,797) and those not in this sample (n = 306,317), as well as the whole available UKB sample (n = 385,163), because the accelerometer-derived sub-sample were recruited non-randomly. In addition, we compared the 1SMR estimates of self-reported sleep traits (sleep duration, chronotype, insomnia symptoms) on HbA1c/glucose in this study (n = 73,797) with those 1SMR estimates, previously published in nearly all UKB participants(14) (n = 336,999, White British ancestry). Similar estimates would suggest limited risk of selection bias.

#### Genetic correlation between sleep traits

We used linkage disequilibrium score regression (LDSC)(20) (**Supplementary Information**), as an additional analysis, to aid the interpretation of the MR using accelerometer-derived results and interpret any differences that might be observed between our accelerometer-derived data generated and our previously reported MR effects of self-reported sleep traits on HbA1c/glucose.(14) We assessed genetic correlations between all accelerometer-derived and self-reported sleep traits. For completeness, we also explored genetic correlations of each accelerometer-derived and self-reported traits with HbA1c and glucose. The full summary statistics of all sleep traits were obtained from the Sleep Disorder Knowledge Portal https://sleep.hugeamp.org/. Those for HbA1c and glucose were from the MAGIC consortium.(19)

Whenever we observed strong genetic correlation between any two accelerometer-derived sleep traits (i.e., ≥ 0.7) regarding robustness of the univariable MR estimates, we undertook multivariable Mendelian randomization (MVMR)(34) to explore whether we could determine individual accelerometer-derived sleep trait direct effect (**Supplementary Information**).

## Results

### Baseline characteristics

**Figure 1** showed the flow of participants in the UKB sub-sample where the 1SMR analyses were conducted. Participants in the accelerometer-derived sub-sample were more likely to have never smoked, have completed advanced-level education, have a lower prevalence of diabetes and a lower mean BMI than those in either comparison group (i.e., (i) UKB European participants without accelerometer-derived data and (ii) all UKB European participants with available genetic data). Other characteristics, including self-reported sleep traits were similar across the three groups (**Table 1**).

**Table 1.** Characteristics of UK Biobank participants by the status of having accelerometer-derived sleep data.

| Variable | Participants with<br>accelerometer-derived<br>sleep traits<br>Mean (SD) / % | Participants without<br>accelerometer-derived sleep traits *<br>Mean (SD) / % | All available UKB<br>participants with<br>genetic data<br>Mean (SD) / % |
| --- | --- | --- | --- |
| **n** | 73,797 | 306,317 | 385,163 |
| Age (years) | 56.3 (7.8) | 56.8 (8.1) | 56.7 (8.0) |
| Female, Sex, % | 56.0% | 53.6% | 54.0% |
| Body mass index (kg/m<sup>2</sup>) | 26.7 (4.5) | 27.6 (4.8) | 27.4 (4.8) |
| Accelerometer-derived sleep traits |  |  |  |
| Sleep duration, hour | 7.29 (0.86) | - | - |
| Fragmentation / episode, times | 17.2 (3.6) | - | - |
| Efficiency, % | 76% (7%) | - | - |
| L5 timing, hour; time | 27.3 (1.1); 3:18 a.m. | - | - |
| M10 timing, hour; time | 13.7 (1.2); 1:42 p.m. | - | - |
| Self-reported sleep duration | 7.2 (1.0) | 7.2 (1.1) | 7.2 (1.1) |
| Insomnia |  |  |  |
| Never/rarely or sometimes, % | 73% | 71% | 72% |
| Usually, % | 27% | 29% | 28% |
| Chronotype |  |  |  |
| Morning, % | 23% | 24% | 24% |
| More ‘morning’ than ‘evening’, % | 34% | 32% | 32% |
| Do not know, % | 10% | 10% | 10% |
| More ‘evening’ ‘than ‘morning’, % | 25% | 26% | 26% |
| Evening, % | 8% | 8% | 8% |
| HbA1c (mmol/mol) / log(hbA1c) | 35.3 (5.5) / 3.55 (0.14) | 36.1 (6.8) / 3.57 (0.15) | 35.9 (6.5) / 3.57 (0.15) |
| HbA1c ≥ 48 mmol/l, % | 2.3% | 3.7% | 3.4% |
| Glucose (mmol/L) / log(glucose) | 5.1 (1.1) / 1.61 (0.16) | 5.1 (1.2) / 1.62 (0.18) | 5.1 (1.2) / 1.61 (0.17) |
| Diabetes defined by *Eastwood* algorithm, % | 3.2% | 5.0% | 4.6% |
| Diagnosed sleep apnoea §, % | 0.4% | 0.5% | 0.5% |
| *Townsend* deprivation index † | -1.8 (2.8) | -1.4 (3.1) | -1.5 (3.0) |
| Most affluent (the lowest 10%) | 11% | 10% | ref (≤ -4.6) |
| Most deprived (the highest 10%) | 7% | 11% | ref ≥ 3.1) |
| Smoking |  |  |  |
| Never, % | 56.7% | 53.3% | 54.0% |
| Former, % | 36.5% | 35.4% | 35.6% |
| Current, % | 6.8% | 11.3% | 10.4% |
| Alcohol intake ‡ |  |  |  |
| Daily, % | 23.5% | 20.7% | 21.3% |
| One to four times a week, % | 51.6% | 49.6% | 50.0% |
| One to three times a month, % | 10.8% | 11.2% | 11.1% |
| Never / occasionally, % | 14.1% | 18.4% | 17.6% |
| Education (ISCED codes) |  |  |  |
| College or university degree /<br>NVQ or HND or HNC or<br>equivalent, % | 58.3% | 46.4% | 48.8% |
| Other prof.equal. Eg: nursing,<br>teaching, % | 12.7% | 12.1% | 12.2% |
| A levels / AS levels or<br>equivalent, % | 6.3% | 5.5% | 5.7% |
| O levels / GCSEs or equivalent<br>/ CSEs or equivalent, % | 14.6% | 17.2% | 16.7% |
| None of the above, % | 8.1% | 18.8% | 16.6% |
| Days of vigorous physical activity per week ‡ |  |  |  |
| 0 – 1 day, % | 49.9% | 52.1% | 51.6% |
| 2 -3 days, % | 32.4% | 29.0% | 29.7% |
| 4 – 7 days, % | 17.7% | 18.8% | 18.6% |

\* The number of participants without accelerometer-derived data (n = 306,317) is obtained via the whole European sample (n = 385,163) minus the number of raw accelerometer-derived sample (n = 78,846, which had not accounted for activity-monitor data quality check).
† Townsend deprivation index was calculated using data from the preceding national census output areas, where each participant was assigned, a continuous score corresponding to the output area of their postcode location. A higher index indicates a greater level of deprivation.
‡ Alcohol intake was categorized and adjusted as “Daily”, “One or four times a week”, “Once or twice a week”, “One to three times a month”, “Occasionally”, “Never”; vigorous physical activity was categorized as days from 0 to 7 per week. (Details in Supplementary Information)
§ Sleep apnea (ICD-10) was diagnosed from the Hospital Episode Statistics (HES) data.

### MR results

In 1SMR analysis, we generated unweighted allele scores for both accelerometer-derived and self-reported sleep traits as the total number of sleep trait increasing alleles present for each participant, based on SNPs identified in the relevant GWAS. **Supplementary Table S1** provides details of each SNP. The R^2^ explained by the allele scores varied from 0.04% for M10 timing (F-statistic: 30) to 0.74% for sleep fragmentation (F-statistic: 553) among accelerometer-derived traits, and from 0.54% for sleep duration (F-statistic: 401) to 2.12% for chronotype (F-statistic: 1593) among self-reported traits (**Supplementary Table S2**).

In the two sets of 2SMR (i.e., 2SMR-UKB and 2SMR-MAGIC), the R^2^ explained and the F-statistics for sleep traits were similar, ranging from 0.04% for M10 timing (mean F-statistic: 37) to 0.91% for sleep fragmentation (mean F-statistic: 37) among accelerometer-derived traits, and from 0.68% for sleep duration (mean F-statistic: 40) to 2.78% for chronotype (mean F-statistic: 57) among self-reported traits (**Supplementary Table S2**).

1SMR suggested longer mean accelerometer-derived sleep duration reduced mean HbA1c levels (−0.11, 95% CI: −0.22 to 0.01 SD per hour longer over 24-hours). However, the association was attenuated to the null in sensitivity analyses accounting for any possible horizontal pleiotropy (i.e., collider-correlated estimates(32)) in 1SMR; 2SMR main and sensitivity results provided no robust evidence of an effect of accelerometer-derived sleep duration on HbA1c (**Figure 2** and **Supplementary Table S3**). For all other accelerometer-derived sleep traits, MR estimates did not support any evidence of causal effects on HbA1c (**Figure 2** and **Supplementary Table S3**). Results (1SMR and 2SMR-UKB) were broadly consistent when participants with diabetes were excluded (**Supplementary Table S3** and **S4**). There was no evidence suggesting any effect of accelerometer-derived sleep traits on glucose (**Figure 3** and **Supplementary Table S4**).

**Figure 2.**
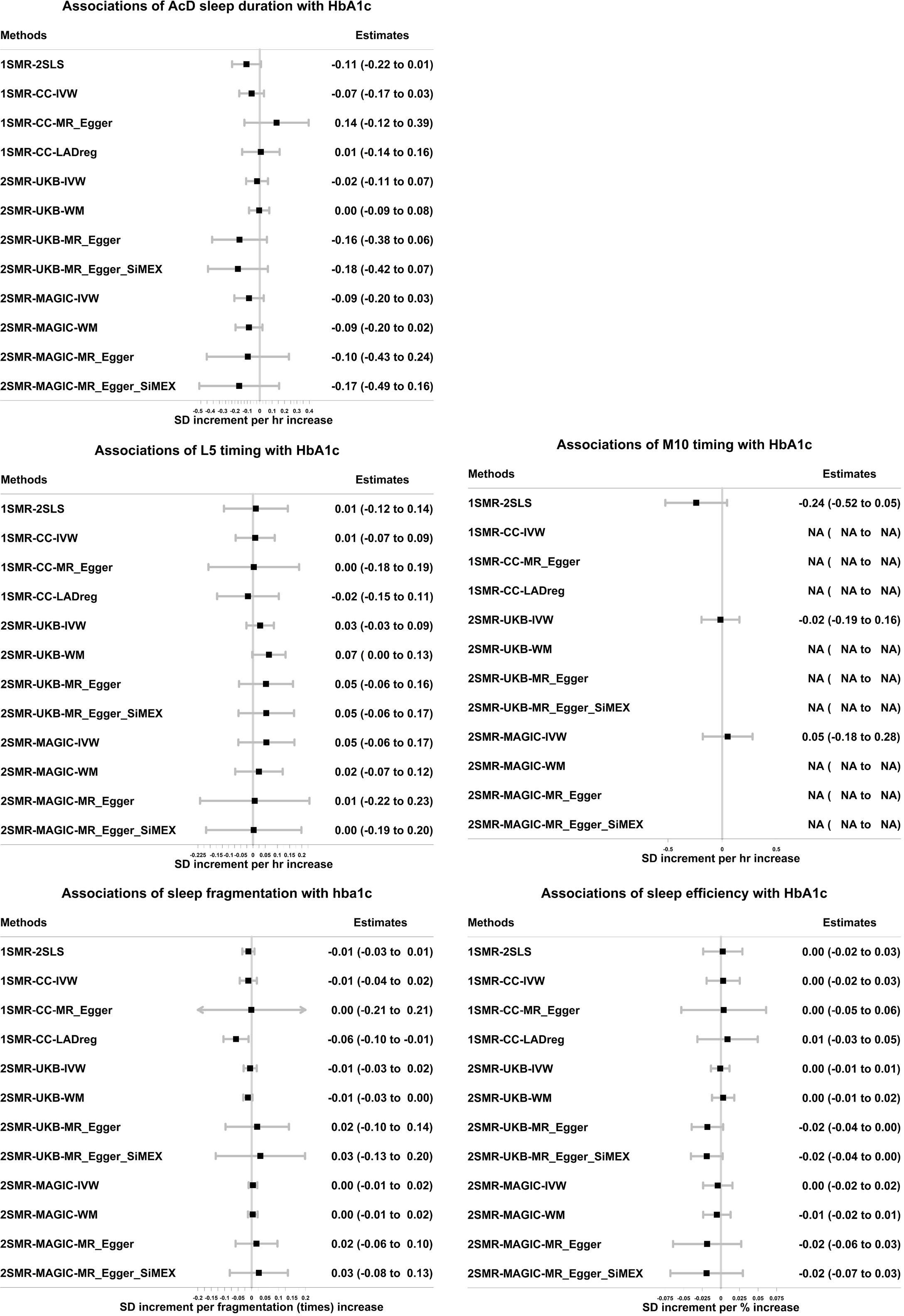
Associations of accelerometer-derived sleep traits with HbA1c in one-sample and two-sample Mendelian randomization. 1SMR-2SLS: one-sample MR with two-stage least square method. 1SMR-CC-IVW, MR_Egger, LADreg: one-sample Mendelian randomization with collider-correction in inverse-variance weighted, MR-Egger, and LAD regression respectively. 2SMR-UKB/MAGIC-IVW, WM, MR-Egger, MR-Egger_SiMEX: two-sample MR with inverse-variance weighted, weighted median, MR-Egger, MR-Egger with simulation extrapolation SiMEX respectively. 1SD HbA1c in the UK Biobank with accelerometer-derived data is 0.14 log mmol/mol; 1SD HbA1c in the sub-sample of UK Biobank without accelerometer-derived data is 0.15 log mmol/mol; 1SD HbA1c in the MAGIC is 0.41% Only 1 SNP predicting M10 timing was identified. As such, the 1SMR-CC was not reliable in the simulation process, and the 2SMR-WM/Egger estimates were not available.

**Figure 3.**
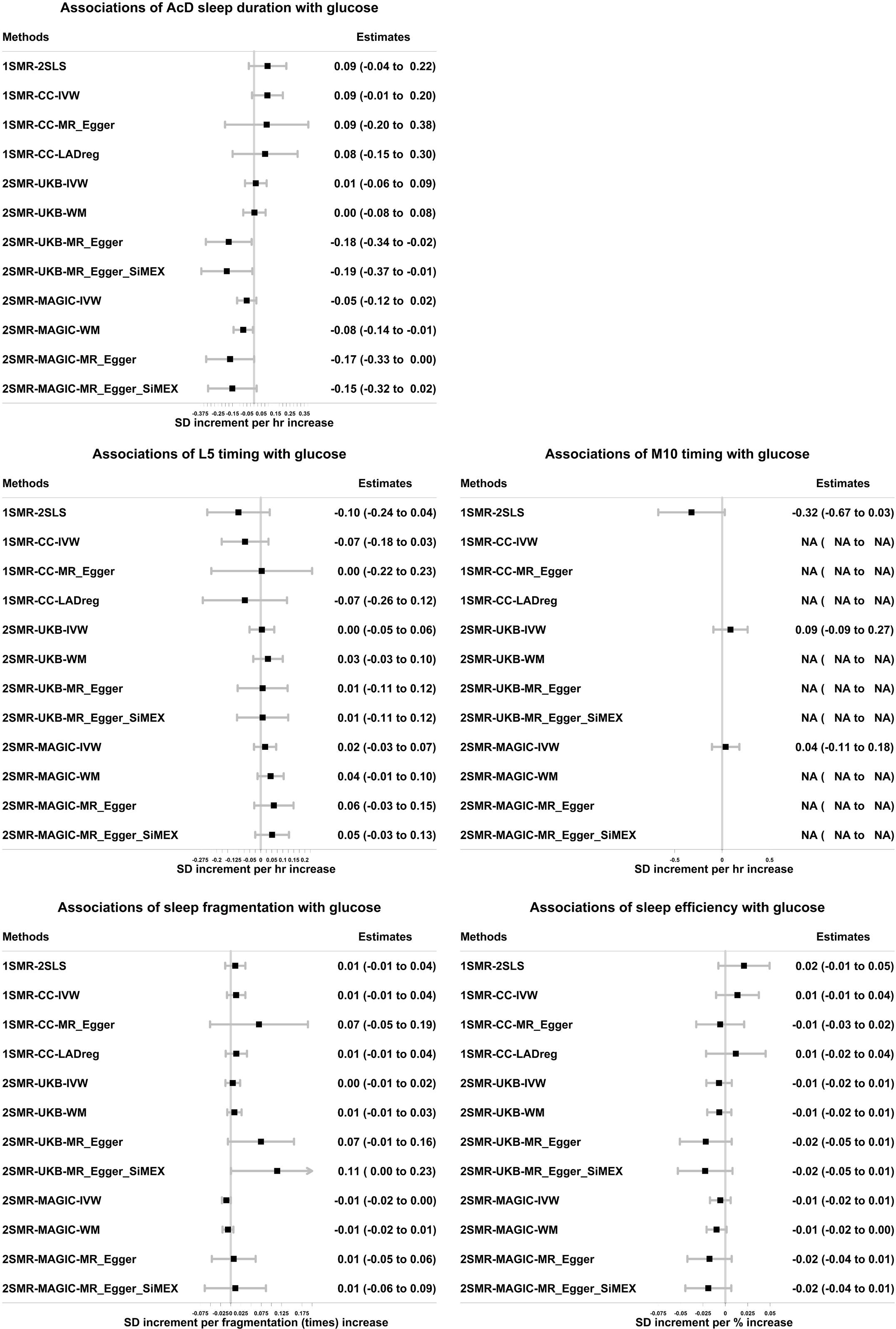
The associations of accelerometer-derived (AcD) sleep duration, mid-point least active 5-hours (L5), mid-point most active 10-hours (M10), sleep fragmentation, and sleep efficiency with glucose in one-sample Mendelian randomization in the UK Biobank and in two-sample Mendelian randomization in UK Biobank (UKB) and MAGIC. 1SMR-2SLS: one-sample MR with two-stage least square method. 1SMR-CC-IVW, MR_Egger, LADreg: one-sample Mendelian randomization with collider-correction in inverse-variance weighted, MR-Egger, and LAD regression respectively. 2SMR-UKB/MAGIC-IVW, WM, MR-Egger, MR-Egger_SiMEX: two-sample MR with inverse-variance weighted, weighted median, MR-Egger, MR-Egger with simulation extrapolation SiMEX respectively. 1SD glucose in the UK Biobank with accelerometer-derived data is 0.15 log mmol/l; 1SD glucose in the sub-sample of UK Biobank without accelerometer-derived data is 0.18 log mmol/l; 1SD glucose in the MAGIC is 0.84 mmol/l Only 1 SNP predicting M10 timing was identified. As such, the 1SMR-CC was not reliable in the simulation process, and the 2SMR-WM/Egger estimates were not available. Non-fasting glucose in the 1SMR and 2SMR-UKB estimates; fasting glucose adjusted for BMI in the 2SMR-MAGIC estimates.

In 1SMR, the associations of self-reported traits with HbA1c/glucose in the UKB sub-sample with accelerometer-derived data were consistent, though with wider confidence intervals, with those we previously published using the larger samples(14) (**Supplementary Figure S1**).

### Genetic correlations and MVMR

Strong genetic correlations were demonstrated between the three sleep timing traits (accelerometer-derived L5 timing, M10 timing, and self-reported chronotype; all R_LDSC_ > 0.8). There was modest genetic correlation between accelerometer-derived and self-reported sleep duration (R_LDSC_ = 0.43) and relatively strong genetic correlation between accelerometer-derived sleep duration and sleep efficiency (R_LDSC_ = 0.72). Genetic correlations of self-reported insomnia with both accelerometer-derived efficiency and fragmentation were weak (both R_LDSC_ < 0.18), with modest correlation between accelerometer-derived sleep fragmentation and sleep efficiency (R_LDSC_ = −0.52). There were weak negative genetic correlations of self-reported sleep duration with HbA1c (R_LDSC_ = −0.07) and glucose (R_LDSC_ = −0.07), and weak positive genetic correlation of insomnia with HbA1c and glucose (R_LDSC_ ≤ 0.1) (**Figure 4** and **Supplementary Table S5**).

**Figure 4.**
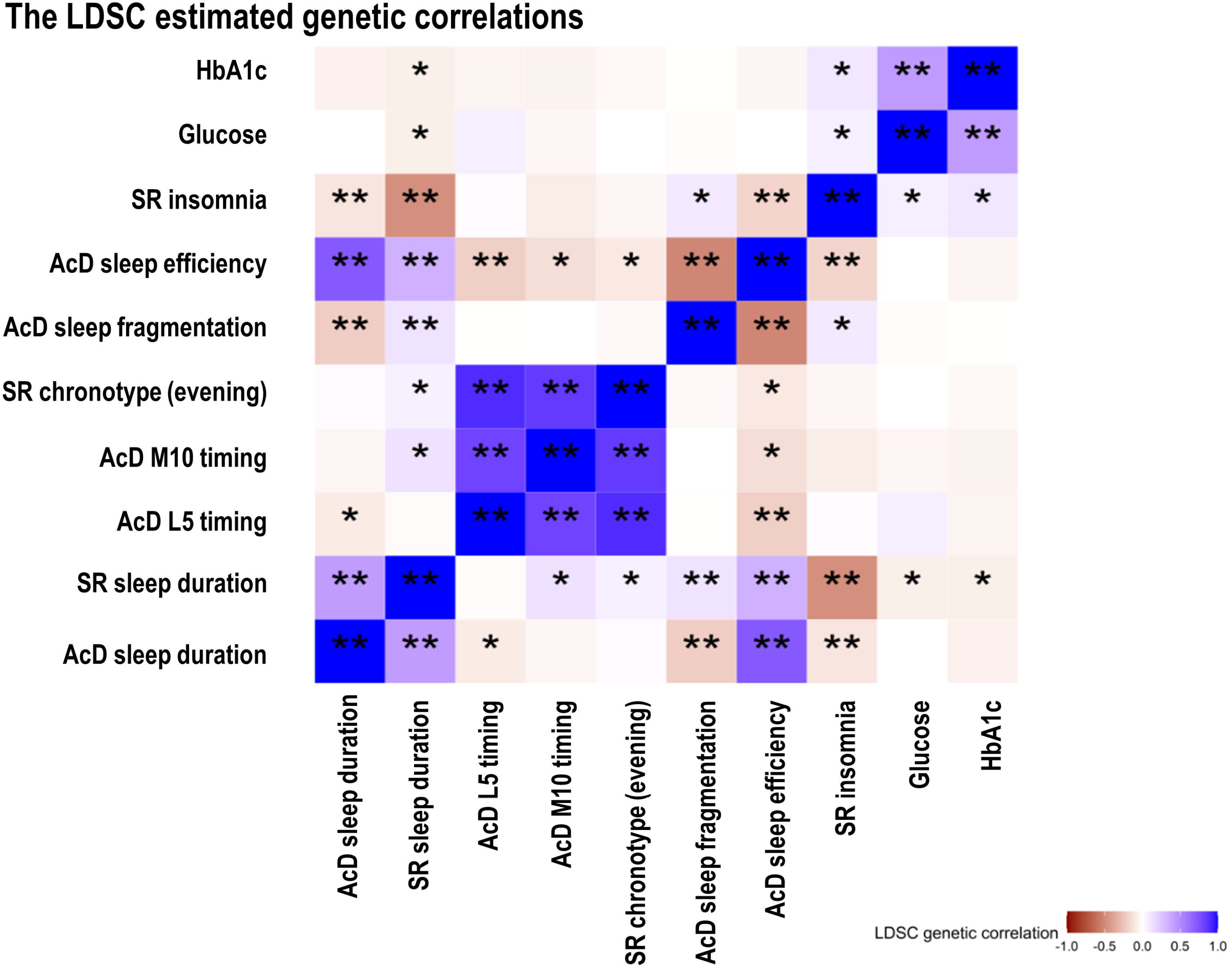
The genetic correlations across accelerometer-derived and self-reported sleep traits and glycaemic traits. * p-value < 0.05 ** p-value < 0.001 The genetic data of glucose was fasting and was BMI-adjusted

We repeated MR analyses with mutual adjustment using MVMR(34) to account for strong correlations between accelerometer-derived sleep traits (i.e., between L5 and M10, and between accelerometer-derived sleep duration and efficiency). These results did not differ from the main results, suggesting no independent causal effect of L5, M10, accelerometer-derived sleep duration or efficiency on HbA1c or glucose (**Supplementary Table S6**).

## Discussion

In this first MR study to explore causal effects of accelerometer-derived sleep traits on glycaemia. We found no robust evidence that any assessed sleep trait causally affected HbA1c or glucose, including across a suite of sensitivity analyses and in MVMR adjusting for between-trait correlations. The null effects of accelerometer-derived sleep traits were unlikely to be explained by selection bias. We showed strong positive genetic correlations between accelerometer-derived L5 and M10 timing, and self-reported chronotype, suggesting that accelerometer-derived and self-reported measures for sleep timing were capturing the same trait. By contrast, positive correlations between accelerometer-derived and self-reported sleep duration were modest. Those between self-reported insomnia and two accelerometer-derived measures (i.e., low sleep efficiency and high sleep fragmentation) that might be expected to relate to insomnia were weak. Lastly, we found no effect of sleep fragmentation or efficiency on HbA1c, though effects of insomnia were identified previously.(14) Accelerometer-derived measures of sleep duration and sleep quality might not simply be ‘objective’ measures of self-reported sleep duration and insomnia, but rather they might capture different underlying sleep characteristics.

Our MR findings do not support the observational associations of accelerometer-derived sleep measures (e.g., shorter sleep duration,(6) lower sleep efficiency,(7) higher sleep fragmentation(8)) with higher glycaemia levels. These observational relationships might be explained by residual confounding, as well as reverse causality as most previous observational studies were cross-sectional. For example, undiagnosed hyperglycaemia might cause nocturia(35) and/or neuropathic pain,(36) which could result in reduced sleep duration and poor sleep quality. Our MR findings also do not support data from randomised controlled trials which have shown that sleep restriction reduces insulin sensitivity, at least in short-term studies.(11).

Sleep characteristics might be captured differently through assessment of self-reported and accelerometer-derived traits. For instance, the self-reported sleep duration question includes naps but this is not the case for accelerometer-derived sleep duration. The genetic correlations (R_LDSC_ = 0.43) also indicated a modest correlation. The null MR estimates of accelerometer-derived sleep fragmentation and efficiency (assumed measures of insomnia(5, 9)) with HbA1c contrasted with previous MR results suggesting that self-reported frequent insomnia symptoms results in higher HbA1c levels.(14–16) Several factors could explain these differences. Self-reported insomnia is by definition experienced, and that experience, rather than the sleep disturbance, might cause or be a proxy for adverse mental or physical health outcomes, such as depression/anxiety,(38) endocrine disorders,(39) and/or appetite changes,(40) that influence HbA1c. Besides, sleep can be disturbed in ways not detectable by actigraphy or even polysomnography. Therefore, accelerometer-derived sleep fragmentation and efficiency might only reflect insomnia status in terms of sleep changes, but not mental or physical changes. The low genetic correlations of accelerometer-derived sleep fragmentation (R_LDSC_ = 0.09) and efficiency (R_LDSC_ = −0.18) with self-reported insomnia supports this idea to some extent. It is also possible that genetic contributions to self-reported and accelerometer-derived measures of insomnia/sleep quality differed, though heritability estimates using UKB data suggested these were similar (17% for self-reported insomnia(41) and 22% for accelerometer-derived fragmentation(18)). Further studies exploring what might contribute to weak/modest correlations between self-reported and accelerometer-derived measures of sleep duration and quality/insomnia are important, though noting that actigraphy data provides limited data about sleep physiology in terms of macro or microstructure.(42) A key strength of this study is its novelty in using MR to explore potential causal effects of accelerometer-derived sleep traits on HbA1c and glucose. We conducted 1SMR, 2SMR, and a range of sensitivity analyses to explore genetic instrument validity. The consistency of findings across these methods, and across samples, increases confidence in our conclusion that accelerometer-derived sleep traits do not have causal effects on HbA1c or glucose.

We acknowledge the following potential limitations. Our results could be influenced by selection bias,(43) due to the low recruitment into UKB (5.5% participation(21)), as well as the non-random selection of UKB participants into the accelerometer-derived sub-sample resulting in a healthier accelerometer-derived sub-sample of UKB. Whilst the low participation into UKB could result in selection bias,(44) similar observational and MR associations with a range of outcomes have been obtained in meta-analyses with/without UKB participants being included, where other cohorts had higher response rates (i.e. ≥ 70%).(45, 46) Besides, in this study, when we compared 1SMR estimates of self-reported sleep traits on HbA1c/glucose in the accelerometer-derived sub-sample to the same results in a much larger UKB sample that we previously published,(14) we found similar results, suggesting minimal bias due to selection. Results did not differ in sensitivity analyses that excluded participants with diabetes, suggesting our results are not influenced by having diabetes or treatment with hypoglycaemics. We assumed the genetic instrument reflects lifetime exposure. Although the accelerometer-derived data was obtained sometime after the measure of HbA1c, there was unlikely a concern of reverse causality in an MR design. If not the case (i.e., there was reverse causality), we would expect the results to be biased away from the null, which is contrary to our findings. We used genetic variants that passed a p-value threshold of p < 5 x 10^-8^ in UKB, but with limited evidence of replication in an independent cohort.(18) Without further replication in larger studies, it was possible that some of the 47 SNPs were false positives and/or had inflated associations with sleep traits, which could result in both our 1SMR and 2SMR results being biased towards the null.(47) However, a recent study has suggested the use of SNPs from GWAS that have not been independently replicated may not result in notable bias.(48) Participants were predominantly of European ancestry, meaning our findings may not generalise to other ancestries. Lastly, our study assumed linear associations between accelerometer-derived sleep traits and HbA1c/glucose. If there was a symmetrical U-shaped association, this linear assumption would bias results toward the null.

## Conclusions

We found little evidence to support causal effects of any accelerometer-derived sleep trait on HbA1c or glucose levels across a wide range of MR methods. We cannot rule out non-linear (e.g., U-shaped) effects and acknowledge the need for further GWAS and MR studies of accelerometer-derived traits in larger diverse populations.

## Data Availability

All UK Biobank data are available to the research community via an application, with full details of this process provided on the study website (https://www.ukbiobank.ac.uk/enable-your-research/apply-for-access). Summary data on sleep traits and glycaemic traits are available unrestricted and can be downloaded, as we have for this study from the Sleep Disorder Knowledge Portal https://sleep.hugeamp.org/ and the MAGIC from www.magicinvestigators.org respectively. R scripts for the key analyses are available on GitHub at: https://github.com/jamesliu0501/Accelerometer-derived-sleep-traits-on-HbA1c-MR-project-.git. For statistical code relating to the individual level data analysis in UK Biobank, please contact the corresponding author via.

## Acknowledgements

This research has been conducted using the UK Biobank Resource under application 6818. We would like to thank the participants and researchers from the UK Biobank who contributed or collected data. Data on glycaemic traits have been contributed by MAGIC investigators and have been downloaded from www.magicinvestigators.org. We thank Dr. Eleanor Sanderson (MRC/IEU, University of Bristol) for providing statistical support on multivariable Mendelian randomization.

## Data Availability Statement

All UK Biobank data are available to the research community via an application, with full details of this process provided on the study website (https://www.ukbiobank.ac.uk/enable-your-research/apply-for-access). Summary data on sleep traits and glycaemic traits are available unrestricted and can be downloaded, as we have for this study from the Sleep Disorder Knowledge Portal https://sleep.hugeamp.org/ and the MAGIC from www.magicinvestigators.org respectively.

R scripts for the key analyses are available on GitHub at: https://github.com/jamesliu0501/Accelerometer-derived-sleep-traits-on-HbA1c-MR-project-.git. For statistical code relating to the individual level data analysis in UK Biobank, please contact the corresponding author via <u>.</u>

## Authors’ Contributions

J.L. affirms that this manuscript is an honest, accurate, and transparent account of the study being reported, that no important aspects of the study have been omitted, and that any discrepancies from the study as planned (and, if relevant, registered) have been explained. J.L. carried out the data analysis. M.K.R. and D.A.L. conceived the idea for the study. M.K.R., R.A.E., D.W.R., S.G.A., M.J.C., M.N.W., T.M.F., A.R.W., J.B., and D.A.L. obtained funds for the project. D.A.L. and M.K.R. designed the study. J.L. carried out the data analysis. J.L., R.C.R., E.L.A., D.A.L., M.K.R., and J.B. wrote the initial draft, with subsequent input from other authors. All authors made critical revisions to the paper. J.L., R.C.R., E.L.A., D.A.L., and M.K.R. are the guarantors of this work and, as such, had full access to all the data in the study and take responsibility for the integrity of the data and the accuracy of the data analysis.

## Ethics Approval

The UKB has received ethical approval from the U.K. National Health Service National Research Ethics Service (London, U.K.) (ref 11/NW/0382). This manuscript does not contain any personal or medical information about an identifiable individual.

