## Supplementary Figures for "The role of accelerometer-derived sleep traits on glycated haemoglobin and glucose levels: a Mendelian randomization study"

### **Title**

**Supplementary figures**

Supplementary Figure 1 The one-sample Mendelian randomization estimates of self-reported (SR) sleep traits with: a) HbA1c and b) non-fasting glucose in European UKB participants with accelerometer-derived sleep data, and in all available White British UKB participants.

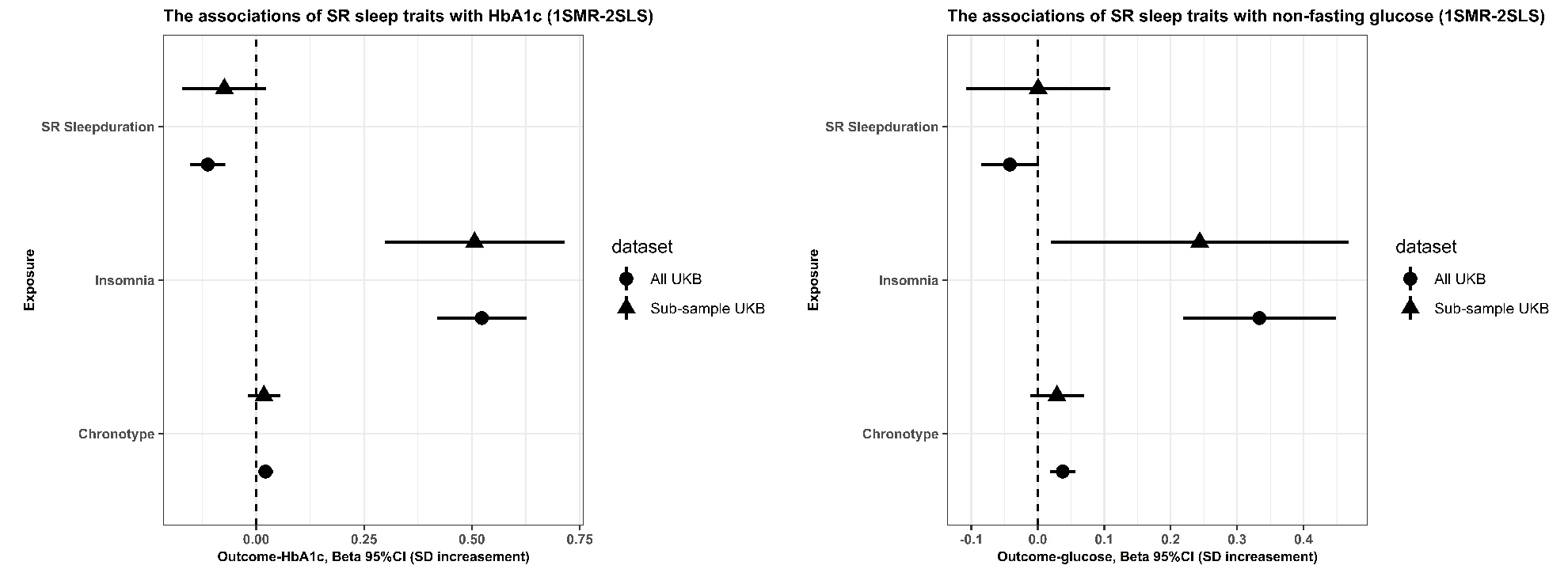

1SMR-2SLS: one-sample MR with two-stage least square method.

SR sleep duration: per hour increase

Insomnia: usually experiencing insomnia symptoms vs sometimes, rarely/never

Chronotype: per category increase to evening preference

1SD HbA1c in the whole UKB is 0.15 log mmol/mol; 1SD HbA1c in the sub-sample of UKB is 0.14 log mmol/mol

1SD glucose in the whole UKB is 0.17 log mmol/l; 1SD glucose in the sub-sample of UKB is 0.15 log mmol/l

Glucose in the UK Biobank was non-fasting.
