## Supplementary Information for "The role of accelerometer-derived sleep traits on glycated haemoglobin and glucose levels: a Mendelian randomization study"

**Title**

* Corresponding author:

Dr. Junxi Liu

### **UK Biobank**

The UK Biobank (UKB) recruited 503,317 participants out of 9.2 million (5·5% response rate) eligible adults, aged between 40 and 69 years, in the UK from 2006 to 2010.(1, 2) At recruitment, a wide range of data were collected for health research after obtaining the participants’ informed consent. Meanwhile, venous blood samples were taken and used to assay germline genotype, from which single-nucleotide polymorphism (SNP) data were collected.(3)

488,377 (97%) participants were successfully genotyped in the UKB; 49,979 using the UK BiLEVE chip and 438,398 using the UKB axiom chip. Pre-imputation quality control, phasing, and imputation of the UKB genetic data have been described.(4) We focused on 464,711 ‘European’ ancestry’ participants who could be linked with the genetic data in the UKB(5) (‘European ancestry’ is as defined by an in-house k-means cluster analysis performed using the first 4 principal components provided by UKB in the statistical software environment R(6)). We excluded 79,460 participants, accounting for overlapping (366 sex mismatch, 626 sex chromosome aneuploidy, 881 outliers in heterozygosity and missing rates, 8 high relatedness, and 77,866 minimal relatedness), based on the quality control procedure undertaken and the derived files produced by the MRC-IEU (University of Bristol), using the full UKB genome wide SNP data (version 3, March 2018).(6) We also excluded 88 participants who had withdrawn (by 22^nd^ Feb, 2022). As such, there were 385,163 (76.5%) available European participants with baseline characteristics as well as self-reported sleep data. Among this available sample, 78,846 (15·7%) participants completed accelerometer sleep data collection, who had worn a triaxial accelerometer device (Axivity AX3) continuously for up to 7 days in an average of 5 years after the baseline assessment (range 2·8 to 8·7 years).(5) We excluded 5,049 individuals, accounting for overlapping, due to failing activity-monitor data quality control based on the accelerometer-derived sleep traits genome-wide association study (GWAS).(5) These included individuals flagged by UKB as having data problems (n = 2,867), suboptimal wear time (n = 1,834, good wear time: at least three days (72 hours) of data and also having data in each one-hour period of the 24-hour cycle (scattered over multiple days)), suboptimal calibration (n = 4, a participant should be excluded from further analysis because recalibration by the preceding or subsequent measurement was not possible due to insufficient data), unable to calibrate activity data on the device worn itself requiring the use of other data (n = 183), and outliers (> 3rd quartile + 1·5 * IQR) (i.e., number of data recording errors (n = 851), interrupted recording periods (n = 1,652), and duration of interrupted recoding periods (n = 1,652)). At the end, 73,797 (14·7%) participants remained in the UKB sub-sample with accelerometer-derived measured sleep traits for the main analyses (**Figure 1**).

#### **Baseline characteristics**

Information on lifestyle and socio-demographic characteristics were obtained using a touchscreen questionnaire at the baseline assessment. Of the lifestyle and environment questions, participants were asked about their smoking status (categorised into ‘never’, ‘former’ or ‘current’) and their alcohol intake frequency (categorised into ‘never’, ‘occasionally’, ‘1-3 times a month’ ‘once or twice a week’, ‘3-4 times a week’ or ‘daily’); Participants were also asked how many days in a typical week that they would do 10 or more minutes of vigorous physical activity (“activities that make you sweat or breathe hard such as fast cycling, aerobic exercise and heavy lifting”); Participants were asked which qualifications they had. A categorial variable was generated for education in the UKB corresponding to 5 International Standard Classification of Education (ISCED) codes based on the years of education in UKB (5: College or university degree / NVQ or HND or HNC or equivalent; 4: Other prof.equal. eg: nursing, teaching; 3: A levels / AS levels or equivalent; 2: O levels / GCSEs or equivalent / CSEs or equivalent; 1: None of the above). Townsend deprivation index(7) was calculated based on the preceding national census output areas, where each participant was assigned a continuous score corresponding to the output area in which their postcode was located. A higher index indicates a greater level of deprivation.

At the initial Assessment Centre visit, height (cm) was measured using a Seca 202 device in all participants in the UKB along with sitting height while weight (kg) was measured by a variety of means, which was amalgamated into a single weight variable. Body mass index (BMI) was calculated from height and weight in kg/m^2^, which were measured at UKB assessment centres (fieldworker assessed weight and height at baseline. Standing height (cm) was measured using a Seca 202 device following a protocol and training. Weight (kg) was measured by a variety of means during the initial Assessment Centre visit. This field amalgamates these values into a single item).

Diagnosed sleep apnoea (ICD-10) was obtained from the Hospital Episode Statistics (HES) data (code G47.33) in the UKB. We ensured that the diagnosis occurred before the baseline UKB assessment using dates of diagnosis and UKB assessment.

#### **Self-reported sleep traits**

Self-reported sleep duration was assessed by asking: “How many hours sleep do you get in every 24 hours? (please include naps)”. The answer could only contain integer values.

self-reported chronotype was assessed in the question “Do you consider yourself to be?” with the following answers: “Definitely a ‘morning’ person”, “More a ‘morning’ than an ‘evening’ person”, “Do not know”, “More an ‘evening’ than a ‘morning’ person”, “Definitely an ‘evening person”, and “Prefer not to answer” which were coded from 1 to 5 (from morning to evening per category increase) and missing respectively.

The UKB assessed self-reported insomnia as insomnia complaints. To assess the frequency of insomnia symptoms, participants were asked: “Do you have trouble falling asleep at night or do you wake up in the middle of the night?” with responses “Never/rarely”, “Sometimes”, “Usually”, “Prefer not to answer”, and “Do not know”. Those who responded “Prefer not to answer” or “Do not know” were set into missing. We derived a binary variable for the frequency of insomnia symptoms where “Usually” was coded as 1 and “Never/rarely” or “Sometimes” were coded as 0.

#### **HbA1c and glucose measurement**

HbA1c was measured in red blood cells by HPLC on a Bio-Rad VARIANT II Turbo analyzer and glucose was assayed in serum by hexokinase analysis on a Beckman Coulter AU5800.(24) Samples were assumed to be non-fasting, because participants were not advised to fast before attending. They were asked to record the last time they ate or drank anything other than water before attending the clinic and those answers were used to derive ‘fasting time’ prior to sampling. During routine quality control checks, the UKB laboratory team observed that, due to a sample processing error, ~ 8% of the glucose assay results were lower than expected and therefore a ‘dilution correction factor’ was provided and applied to these results.(25) The HbA1c samples were not affected. We used HbA1c as our primary outcome because it provides a stable measure over a period of ~ four weeks and is therefore less prone to regression dilution bias and is more statistically efficient compared with non-fasting glucose which we explored as a secondary outcome.

### **Genetic variants**

#### **Genetic variants of accelerometer-derived sleep traits**

The genetic variants associated with the accelerometer-derived sleep traits were obtained from a recent GWAS conducted in a white European subset of UKB (n = 85,670), where 44 genetic associations at genome-wide significant (p < 5 x 10^-8^) were identified (11 for sleep duration, 6 for mid-point least active 5-hours (L5 timing), 1 for mid-point most active 10-hours (M10 timing), 21 for sleep fragmentation, 5 for sleep efficiency) (**Supplementary Table 1** and **2**). This GWAS study imputed 11,977,111 genetic variants using the Haplotype Reference Consortium imputation reference panel with a minimum minor allele frequency (MAF) > 0·1% and imputation quality score (INFO) > 0·3. The genetic associations were obtained using a linear mixed model (LMM) adjusting for the effects of population structure, individual relatedness, age at accelerometer assessment, sex, study centre, season of accelerometer wear, and genotype array.(5)

#### **Genetic variants of self-reported sleep traits**

For self-reported sleep duration, 78 SNPs (78 loci) for self-reported sleep duration (p < 5 × 10^−8^) were reported in a GWAS analysis in 446,118 adults of European ancestry (**Supplementary Table 1** and **2**). BOLT-LMM and an additive genetic model (adjusting for age, sex, 10 principal components of ancestry, genotyping array, and genetic correlation matrix with a maximum per SNP missingness of 10% and per sample missingness of 40%) were used to perform GWAS analysis. It has a hard-call genotype threshold of 0.1, SNP imputation quality threshold of 0.80, and a minor allele frequency (MAF) threshold of 0·001. Genetic association analysis for the X chromosome was performed using the genotyped markers on the X chromosome with the additional sex flag in PLINK.(8) 77 SNPs with individual data were identified in the UKB (rs1776776 was not available), which were applied in all the analyses throughout the study. The summary statistics can also be extracted from the Sleep Disorder Knowledge Portal <http://www.kp4cd.org/dataset_downloads/sleep>.

For self-reported chronotype (morning chronotype was reported in the discovery GWAS, but we converted it to evening chronotype in the corresponding MR analyses), 351 SNPs (351 loci) for chronotype (p < 5 × 10^−8^) were identified in a GWAS analysis with 697,828 European participants from UK Biobank (n = 451,454) and 23andMe (n = 248,100) participants (**Supplementary Table 1** and **2**). In the UKB. the genome-wide associations were obtained using BOLT-LMM adjusting for population structure, individual relatedness, age sex, study centre, and genotyping release. In the 23andMe morning person GWAS, the summary statistics were generated via logistic regression (an additive model) adjusting for age, sex, the first four principal components, and a categorical variable representing genotyping platform. A meta-analysis was performed using the results from the UKB chronotype GWAS and the 23andMe morning person GWAS, therefore obtaining a large sample size and high statistical power.(9)

For self-reported insomnia, 248 SNPs (202 loci) for insomnia symptoms (p < 5 × 10^−8^) were identified in a meta-analysis of GWAS with 1,331,010 European participants from UK Biobank (n = 386,533) and 23andMe (n = 944,477) participants (**Supplementary Table 1** and **2**). GWAS on insomnia in UKB was performed in PLINK, using logistic regression adjusting for age, sex, genotype array, and 10 genetic principal components. In the 23andMe, association testing for each SNP was conducted using logistic regression adjusting for age, sex, and the top 5 principal components.(10)

### **Statistical analyses**

#### **One-sample Mendelian randomization (1SMR)**

*Two-stage least squares instrumental variable analyses*

We identified SNPs in the UKB data that were aligning with the genome-wide significant (p < 5 x 10^-8^) SNPs found in the discovery accelerometer-derived (5) and self-reported (8-10) sleep traits GWAS (i.e., the direction of specific sleep traits’ increasing allele). For self-reported chronotype, we aligned evening preference alleles for a better interpretation, where ‘definitely a morning person’ is the reference category with ordering from this category to more ‘eveningness’ categories. We then extracted the genetic variants from the UKB Haplotype Reference Consortium reference panel dataset. These data have undergone extensive quality control checks including removal of related participants (third degree or closer) and non-White British participants based on questionnaire and PCA.(11) As such, for accelerometer-derived sleep traits, 11 SNPs genetically predicted accelerometer-derived sleep duration; 6 SNPs genetically predicted L5 timing, 1 SNP genetically predicted M10 timing, 22 SNPs genetically predicted sleep fragmentation, 5 SNPs genetically predicted sleep efficiency; for self-reported sleep traits, 77 SNPs genetically predicted self-reported sleep duration, 245 SNPs genetically predicted insomnia symptoms, and 341 SNPs genetically predicted chronotype. Lastly, the unweighted allele scores of each sleep trait were generated by summing the number of effect alleles harboured by each individual, which, when unweighted, suffer from less bias due to sample overlap (i.e., the samples used for performing a discovery GWAS overlap with that used to perform MR).(12) Two-stage least squares (2SLS) instrumental variable analyses were performed with adjustment for assessment centre and 40 genetic principal components to minimize confounding by population stratification,(13) as well as baseline age, sex and genotyping chip, fasting time and dilution factor (for glucose only) to reduce random variation. The F-statistic and variance explained (R^2^) of the unweighted allele score were calculated via a linear model (i.e., sleep trait ~ unweighted allele score).

*Collider-correction method*

The method termed `collider-correction’ enables weak instrument and pleiotropy robust 2SMR methods to be applied to one-sample data to obtain causal estimates.(14) Naively apply summary data MR methods to the one-sample context would result potentially anti-conservative weak instrument bias due to correlated error. The collider-correction method is based on a generalization of the algorithm described in *Dudridge* et al , to adjust for collider bias in genetic association studies of disease progression.(15) This method artificially induces and then corrects for collider bias, with an additional simulation extrapolation (SiMEX) correction(16) step to cope with weak instrument bias. Further methodological details are provided in the link publication(14) (<https://journals.plos.org/plosgenetics/article?id=10.1371/journal.pgen.1009703>). Applying the combination of collider-correction and two-sample Mendelian randomization (2SMR) methods as a sensitivity analysis, it provides an alternative to account for both pleiotropy and weak instrument bias in a 1SMR setting, which is a less biased but more precise causal estimate comparing with the application of standard 2SMR methods. Correspondingly, the SiMEX would adjust for the measurement error of the SNP-exposure association which causes ‘dilution bias’ in 2SMR. Besides, the 1SMR setting guarantees the ‘two cohorts’ are homogeneous (the SNP-exposure and SNP-outcome associations are from the same cohort) as well as accounts for shared subjects across the ‘two cohorts’ (i.e., the independent assumption). Lastly, consistent covariates, as well as model specific covariates, can be adjusted for to obtain the SNP-exposure and SNP-outcome associations.

#### **Two-sample Mendelian randomization (2SMR)**

We conducted 2SMR analyses using the “*TwoSampleMR*” package in R (version 0.4·26).(17) All of the SNPs used to instrument the sleep traits were found to be conditionally independent in the GWAS studies. As such we did not apply the LD clumping function, in order to retain the maximum number of SNPs. If a SNP was unavailable in the outcome GWAS summary statistics, we identified a proxy in strong linkage disequilibrium (LD) with the missing SNP (r^2^ > 0·8). We then performed harmonization of the direction of effects between SNPs in the exposure and outcome GWAS. Palindromic SNPs were harmonized if they were aligned and the minor allele frequency was < 0.3, otherwise they were excluded (This was not applied in the 2SMR-UKB analyses, where we assumed all alleles were presented on the same strand because the SNP-exposure and SNP-outcome associations are both from the UKB).

In primary analysis, we used the inverse-variance weighted (IVW) regression of the Wald ratio for each SNP under a multiplicative random-effects model(18) to obtain estimates for causal effects of the sleep traits on HbA1c and glucose. For the estimates of self-reported insomnia symptom, we converted the results from the multiplicative log odds scale to a difference in risk scale by $\beta=\log OR* \mu*\left( 1-\mu\right), se= {se}_{\log OR}* \mu*\left( 1-\mu\right),$ with $\mu= {n_{case}}/{(n_{case}+ n_{control})}$(19) to enable the comparison of the 2SMR estimates to the 1SMR results.

The mean F-statistic in the 2SMR was equal to the mean of the individual F-statistic of each SNP (individual F statistic was equal to SNP specific estimate: BetaXGi^^2^ / seXGi^^2^ (BetaXGi and seXGi were obtained from the discovery GWAS)). The R^2^ was calculated via R^2^ = K * F / (N -K - 1 + K*F) (F is the mean F-statistic in above, K is equal to the number of SNP applied, N= sample size in the discovery GWAS).

#### **Cross-trait linkage disequilibrium score regression (LDSC)**

This method relies on the fact that the GWAS effect size of a given SNP incorporates the effects of all SNPs in linkage disequilibrium (LD), which only requires GWAS summary statistics to estimate the genetic correlations regardless of sample overlap.(20-22) The cross-trait LDSC equation is

$$E\left[ z_{1j}z_{2j}|\mathcal{l}_{j} \right]=\frac{\sqrt{N_{1}N_{2}} \varrho_{g}}{M}\mathcal{l}_{j}+\frac{\varrho N_{s}}{\sqrt{N_{1}N_{2}}}$$

Where $z_{ij}$ is the z-score for study $i$ and SNP $j$, $N_{i}$ is the sample size for study $i$, $\varrho_{g}$ is the genetic covariance, $\mathcal{l}_{j}$ is the LD Score (computed using 1000 Genomes European data and is appropriate for use with European GWAS data), $N_{s}$ is the number of individuals included in both studies and $\varrho$ is the phenotypic correlation among the $N_{s}$ overlapping samples. The genetic covariance is estimated by regressing $z_{1j}z_{2j}$ against $\mathcal{l}_{j}\sqrt{N_{1j}N_{2j}}$ ($N_{ij}$is the sample size for SNP $j$ in study $i$) and then multiplying the result slope by $M$ (the number of SNPs in the reference panel with minor allele frequency (MAF) between 5 -50%. To obtain a less biased estimate (i.e., bias from variable imputation quality, which is correlated with LD score, and low imputation quality yields lower test statistics), the *ldsc* function (*mung_sumstats.py*) would convert the summary statistics, which would 1) filter to INFO > 0·9 (for studies that provide a measure of imputation quality); 2) filter to sample of MAF above 1% (for studies that provide sample MAF); 3) filter to SNPs in the HapMap 3 panel(23) with a 1000 Genome Project EUR (European) MAF above 5% (for studies that do not provide a measure of imputation quality); 4) remove SNPs with an effective sample size less than 0·67 times the 90^th^ percentile of the sample size (if the sample size varies from SNP to SNP); 5) remove SNPs with a sample size above the maximum GWAS sample size (for meta-analyses with specialty chips (e.g., Metabochip)); 6) remove indel and structural variants; 7) remove strand-ambiguous SNPs; 8) remove SNPs whose alleles do not match the alleles in the 1000 Genomes Project.(22) A more detailed explanation can be found elsewhere(22) and the *ldsc* software and the practical tutorial can be checked <https://github.com/bulik/ldsc/wiki/Heritability-and-Genetic-Correlation>.

#### **Multivariable Mendelian randomization (MVMR)**

Whenever we observed strong genetic correlation between any two accelerometer-derived sleep traits (i.e., ≥ 0·7), we undertook MVMR.(24) In 1SMR setting, we obtained the Sanderson-Windmeijer F statistic(24) of each unweight allele score to assess the instrumental joint strength. In 2SMR setting, we would present and assess conditional F statistic, where the covariance of the conditional F statistic was approximated via using the phenotypic correlation and summary data (i.e., the standard error of the SNP – exposure association). It is a reasonable approximation if the relevant covariance when each SNP only explains a small proportion of each exposure, when the data used to obtain the covariance matches to that used to obtain the SNP - exposure associaton.(25) A conditional F-statistic larger than the rule-of-thumb value of 10 can be considered adequately strong for the purpose of MVMR. To assess potential horizontal pleiotropy in MVMR-2SMR, we calculated the Heterogeneity Q statistics ($Q_{A}$), where rejection of the null indicates one or more variants may be pleiotropic.(25) The conditional F-statistics and Q statistic p-values in 2SMR setting were obtained accounting for the phenotypic correlation between the two exposures.(25) We further presented MVMR-WM and MVMR-MR-Egger estimates accounting for potential unbalanced horizontal pleiotropy. The 2SMR-MVMR analyses were conducted with the “*WSpiller/MVMR: MVMR” R package (*[*https://rdrr.io/github/WSpiller/MVMR/*](https://rdrr.io/github/WSpiller/MVMR/)*).*
